# KNOWLEDGE, PREVALENCE AND RISK FACTORS OF EATING DISORDERS AND PEPTIC ULCER DISEASE AMONG CLINICAL STUDENTS AT THE UNIVERSITY OF NIGERIA TEACHING HOSPITAL, ITUKU-OZALLA, ENUGU

**DOI:** 10.1101/2024.12.01.24318266

**Authors:** Obayi Angelica Chinecherem, Uka Chijioke Ibiam, Ukoha Chidiebere, Ugwunna Nwachukwu Chinedu

## Abstract

**Background:** Eating disorders and peptic ulcer disease are major health problems among young people, of which clinical students are a composite. It is important to study the prevalence, risk factors and knowledge of eating disorders and peptic ulcer disease among clinical students because their characteristics such as skipping meals, high academic workload, social behaviours, etc. make them prone to developing these illnesses.

**Objective:** To determine the prevalence, risk factors and knowledge of eating disorders and peptic ulcer disease among clinical students at the University of Nigeria Teaching Hospital, Ituku-Ozalla, Enugu state.

**Materials and Methods:** A cross-sectional study of 400 clinical students at the University of Nigeria who were selected using multi-stage sampling. A structured and validated questionnaire was administered by the interviewers who also measured the weights and heights of respondents. Statistical Package for the Social Sciences (SPSS) was used to analyse the data. The significance of association was tested with Pearson chi-square, p <0.05 was considered significant.

**Results:** The study showed a low risk and prevalence rate (2.9%) of eating disorders. 95.5% had a mild risk of being diagnosed with PUD and most were knowledgeable about the disease entities. A statistically significant relationship exists between average income of participants **(p=0.018)** and skipping meals **(p=0.023).** There was also a statistically significant relationship between peptic ulcer disease and smoking **(p=0.000)**, consumption of alcohol **(p=0.036)** and herbal concoctions **(p=0.036)**.

**Conclusions:** This study has found a low prevalence, low risk and good knowledge of eating disorders and peptic ulcer disease among clinical students at the University of Nigeria Teaching Hospital, Ituku-Ozalla.

## BACKGROUND

Eating disorders are a broad group of psychological disorders with non-typical eating behaviours leading to physiological effects from overeating or insufficient food intake. ^1^ Some examples of eating disorders are: anorexia nervosa, an obsessive desire to lose weight by refusing to eat, despite being underweight; bulimia nervosa, a disorder characterised by periods of extreme overeating followed by purging, sometimes through self-induced vomiting, fasting, or using laxatives and diet pills; binge-eating, which is out-of-control eating; pica, an abnormal craving for non-food substances; rumination disorder, repetitive regurgitation of food; avoidant/restrictive food intake disorder: a lack of interest in feeding caused by aversive effects of feeding and other specified feeding or eating disorders (OSFED). In the United States of America, about 30 million of all ages and genders suffer from an eating disorder, and of all mental illnesses, eating disorders have the highest mortality rate. ^1^ Eating disorders can cause heart and kidney problems and even death; therefore, early diagnosis and management is important. Treatment modalities for eating disorders are monitoring, mental health therapy, nutritional counselling and sometimes, pharmacotherapy. ^2^

Peptic ulcer disease (PUD) is a common ailment of the alimentary system. ^3^ Peptic ulcers are defects in the gastric or duodenal mucosa that extend through the muscularis mucosa.^4^ PUD is clinically described as a disruption of the continuity of the gastrointestinal mucosal lining which appear as sores of at least 0.5cm in diameter in endoscopic studies. ^5^ It affects about 4 million of the world’s population annually, with incidence of complications in approximately 10–20%. ^5–7^ The principal causes of peptic ulcer disease are *Helicobacter pylori* and/or long-term use of nonsteroidal anti-inflammatory drugs. Studies have identified modifiable risk factors of PUD which include the use of corticosteroids, anticoagulants, coffee, alcohol, smoking, stress, spicy foods, use of unclean water sources and fasting; and non-modifiable risk factors include genetics, age, gender and past history of peptic ulcer disease (PUD). ^8–12^ Epigastric pain is the most common symptom of both gastric and duodenal ulcers. Other symptoms of peptic ulcer disease are dyspepsia (including belching, bloating, distention, and fatty food intolerance), heartburn, chest discomfort, hematemesis or melena resulting from gastrointestinal bleeding, haematochezia, feeling of fullness, belching, etc. ^7^ Complications of peptic ulcer disease include internal bleeding, gut perforation, intestinal obstruction, gastric cancer, etc.

Peptic ulcer disease and eating disorders negatively impact the health and quality of life of affected individuals. A significant number of host communities of higher institutions in Nigeria where most students live, do not have a clean source of water and because unclean source of water (specifically, sewage-contaminated water) is a risk factor of PUD, prevalence rate may rise. ^3^ High prevalence of social media use, peer pressure, body consciousness, and extensive academic workload among clinical students predispose them to eating disorders. Clinical students are at the cusp of becoming healthcare professionals and are the future of the health system. Their knowledge of peptic ulcer disease and eating disorders, its prevalence among them and the risk factors of both conditions will not only affect their own health and quality of life but also that of the larger population who will depend on them for healthcare service delivery.

## METHODOLOGY

The study was carried out in the Enugu and Ituku-Ozalla campuses of the University of Nigeria, the first fully fledged indigenous and autonomous university in Nigeria. A cross sectional descriptive study of undergraduate clinical students at the University of Nigeria, Enugu and Ituku-Ozalla Campuses, Enugu State. The study was carried out among clinical students at the University of Nigeria, Ituku-Ozalla. These include 4th – 6th year medical students, 4th – 6^th^ year dental students and school of nursing students.

All clinical undergraduate students at the University of Nigeria, Ituku-Ozalla who gave consent for the study were included while undergraduate clinical students who were too ill to participate in the study and those who were not on campus during our data collection were excluded from the study.

The minimum sample size for this study was determined by this equation. ^13^

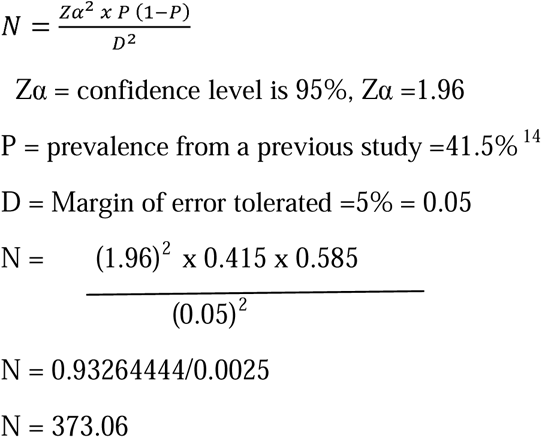

An addition of 10% to make up for attrition will bring the sample size to;

10% of 373.06 = 37.31

Sample size = 373.06 + 37.31 = 410.37

Sample size = 410.37. A sample of 410 will be used.

The number of respondents from each department was determined by multi-stage sampling which involved expressing the number of clinical students in each department as a percentage proportion of the sample size. This ensured that each department was proportionately represented in the study population. Then, three departments: medicine and surgery, dentistry and school of nursing were randomly selected. The questionnaire was then administered to respondents from each department selected by simple random sampling after stratifying them by class and sex to get a representative sample.

A semi-structured, self-administered questionnaire was used for data collection. Some of the questions for eating disorders were adapted from the Eating Attitudes Test (EAT-26) and SCOFF questionnaires while the others were validated by a pilot study. It consisted of five sections. Section A collected sociodemographic data, section B collected data on the respondents’ knowledge of eating disorders, section C collected data on the respondent’s risk factors for eating disorders, section D collected data on the respondents’ knowledge of peptic ulcer disease and, section E collected data on the respondents’ risk factors for peptic ulcer disease.

The heights of respondents were measured with a measuring tape while a weighing scale was used to measure their weights and their body mass index (BMI) calculated using a calculator. The questionnaire was pretested among 50 randomly selected clinical students at Enugu State University of Science and Technology (ESUT) and pharmacy students of University of Nigeria Nsukka to ensure that there were no ambiguous questions.

A score of ‘1’ was assigned to correct response to variables assessing knowledge of PUD in table 5 above. A total of 13 variables were scored, giving a total score of 13. Participants with scores ‘0 – 6’ has poor knowledge of PUD, while participants with score ‘7 – 13’ had good knowledge of PUD. Risk assessment score for peptic ulcer disease was calculated based on positive responses to these questions: people that always/often used NSAIDs longer than 6 months, people that often/always took those plenty things we listed (chocolate, peppery food, etc.), positive family history of peptic ulcer disease, people that take herbal concoction, people that always/often skipped meals, people that take always/often take alcohol, people that often/always smoke, people that have been diagnosed with peptic ulcer disease before, blood group O, more than or equal to 4 in a room (overcrowding), sewage disposal: pit toilet and/or open defecation, source of drinking water is rainfall. A total score of 12 is obtained which is subdivided into three categories: mild risk factors “0 – 4”, moderate risk factors “5– 8”, high risk factors “9 – 12”.

For eating disorders, a score of ‘1’ and ‘2’ were assigned to ‘yes’ and ‘no’ responses respectively to variables that assessed risk factors amongst participants. A total score of 31 was obtained. A risk assessment score was developed for eating disorders using the SCOFF and EAT-26 tests, for which scores between 0-24 and 25-31 were low and high respectively.

Data was analysed with the Statistical Package for the Social Sciences (SPSS) version 26.0 and results were presented in tables, graphs, and pie charts. Summary statistics such as mean, frequency and proportion were used to represent quantitative and qualitative data. Statistical tests were used to test any relationship between variables. Chi-square test was employed to test for a relationship amongst factors that affect prevalence, risk factors and knowledge of eating disorders and peptic ulcer disease.

The research proposal was submitted to the Health Research and Ethics Committee, University of Nigeria Teaching Hospital (UNTH), Ituku-Ozalla, Enugu State and ethical clearance was obtained. During data collection, informed consent was obtained verbally, before administering the questionnaire, and in written form from the participants. Participants were informed that they were free to exit at any point during the study without any consequences.

Out of 410 questionnaires that were distributed, 400 were properly filled and returned.

Limitations of the study: eating disorders and peptic ulcer disease are sensitive topics amongst students and therefore, there is a possibility that some students were not outrightly honest with their responses. However, we allayed any fears by assuring them that their personal information will be used solely for the purposes of this research and that participants were randomly selected by the investigators. Anonymity and confidentiality were maintained.

## RESULTS

### SOCIO-DEMOGRAPHIC CHARACTERISTICS OF PARTICIPANTS

**Table 1.** shows the socio-demographic characteristics of participants. Majority, 309 (77.3%), were within ages 21 – 25 years.

| Variable |  | Frequency | Percentage |
| --- | --- | --- | --- |
| Age (in years) | 16 – 20 | 26 | 6.5 |
|  | 21 – 25 | 309 | 77.3 |
|  | 26 – 30 | 65 | 16.3 |
|  | Total | 400 | 100.0 |
|  | Mean | 23.5 | SD = 2.4 |
| Gender | Male | 200 | 53.1 |
|  | Female | 177 | 46.9 |
|  | Total | 377 | 100.0 |
| Religion | Christianity | 386 | 98.7 |
|  | Islam | 4 | 1.0 |
|  | African Traditional Religion | 1 | .3 |
|  | Total | 391 | 100.0 |
| Ethnicity | Igbo | 368 | 98.4 |
|  | Hausa | 2 | .5 |
|  | Yoruba | 4 | 1.1 |
|  | Total | 374 | 100.0 |
| Marital Status | Single | 386 | 96.7 |
|  | Married | 12 | 3.0 |
|  | Divorced | 1 | .3 |
|  | Total | 399 | 100.0 |
| Department of study | Medicine and Surgery | 304 | 76.0 |
|  | Dentistry | 67 | 16.8 |
|  | School of Nursing | 27 | 6.8 |
|  | Medical Laboratory Science | 2 | 0.5 |
|  | Total | 400 | 100.0 |
| Level of study | 100 – 200 | 9 | 2.3 |
|  | 300 – 400 | 129 | 32.3 |
|  | 500 – 600 | 262 | 65.5 |
|  | Total | 400 | 100.0 |

SD = Standard deviation

**Figure 1:**
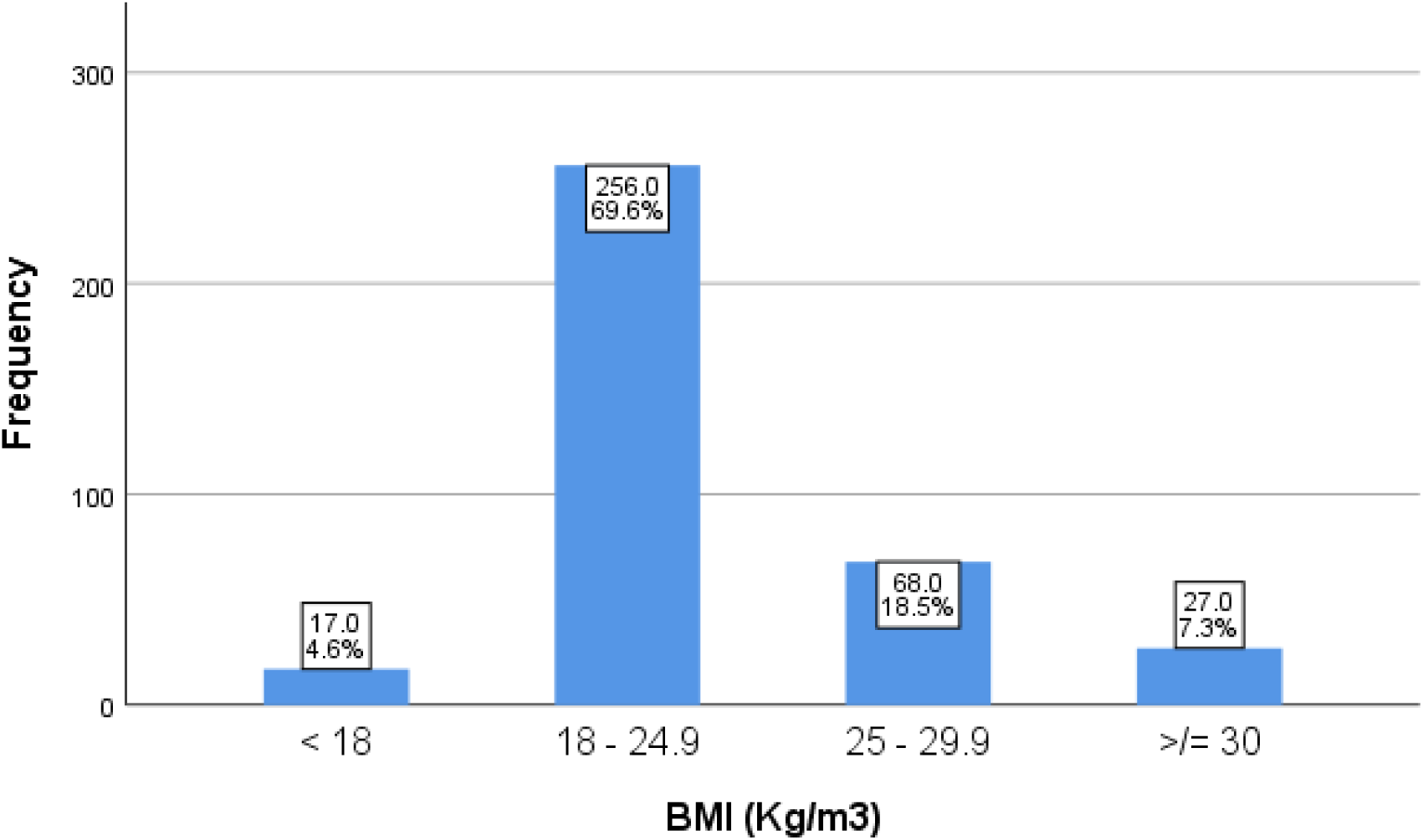
A Simple Bar Chart showing the distribution of BMI amongst participants (N = 368) Most participants (69.6%) had a normal weight (BMI between 18 – 24.9 kg/m^3^).

### SOCIAL HISTORY OF PARTICIPANTS

**Table 2.** shows the social history of participants. The minority of participants take alcohol and smoke – 44.3% and 7.8%.

| Variable |  | Frequency | Percentage |
| --- | --- | --- | --- |
| **Do you take alcohol?** | **Yes** | 177 | 44.3 |
|  | **No** | 223 | 55.8 |
|  | **Total** | 400 | 100.0 |
| **If yes, how often do you take alcohol?** | **Everyday** | 2 | 1.1 |
|  | **Every alternate day** | 0 | .0 |
|  | **2 - 3 times in a week** | 5 | 2.9 |
|  | **Once a week** | 15 | 8.6 |
|  | **once or twice a month** | 20 | 11.5 |
|  | **Occasionally** | 132 | 75.9 |
|  | **Total** | 174 | 100.0 |
| **For how long have you been taking alcohol?** | **less than 6 months** | 13 | 7.6 |
|  | **6 months to one year** | 19 | 11.2 |
|  | **2 - 5 years** | 48 | 28.2 |
|  | **more than 5 years** | 90 | 52.9 |
|  | **Total** | 170 | 100.0 |
| **Do you smoke?** | **Yes** | 31 | 7.8 |
|  | **No** | 369 | 92.3 |
|  | **Total** | 400 | 100.0 |
| **If yes, how often do you smoke?** | **Everyday** | 1 | 3.3 |
|  | **Every alternate day** | 0 | .0 |
|  | **2 - 3 times in a week** | 6 | 20.0 |
|  | **Once a week** | 2 | 6.7 |
|  | **once or twice a month** | 4 | 13.3 |
|  | **Occasionally** | 17 | 56.7 |
|  | **Total** | 30 | 100.0 |
| **For how long have you been smoking?** | **less than 6 months** | 2 | 6.9 |
|  | **6 months to one year** | 5 | 17.2 |
|  | **2 - 5 years** | 17 | 58.6 |
|  | **more than 5 years** | 5 | 17.2 |
|  | **Total** | 29 | 100.0 |

### THE SOCIO-ECONOMIC SITUATION OF PARTICIPANTS

**Table 3.** shows the socio-economic situation of participants with majority (53.3%) receiving an average monthly income between 20,000 and 49,000 naira while minority (6.4%) staying off-campus.

| Variable |  | Frequency | Percentage |
| --- | --- | --- | --- |
| Where do you live? | Hostel | 368 | 93.6 |
|  | Off-campus | 25 | 6.4 |
|  | Total | 393 | 100.0 |
| How many persons are you in a room? | 1 – 2 | 257 | 66.4 |
|  | 3 – 4 | 93 | 24.0 |
|  | More than 4 | 37 | 9.6 |
|  | Total | 387 | 100.0 |
| What is your source of drinking water? | Borehole | 15 | 3.8 |
|  | Sachet water | 375 | 94.0 |
|  | Bottled water | 9 | 2.3 |
|  | Total | 399 | 100.0 |
| What is your method of sewage disposal? | water closet | 358 | 91.1 |
|  | Open defecation | 2 | .5 |
|  | bucket toilet | 33 | 8.4 |
|  | Total | 393 | 100.0 |
| Who is your sponsor? | Parent | 356 | 89.7 |
|  | Siblings | 20 | 5.0 |
|  | Spouse | 8 | 2.0 |
|  | Self | 7 | 1.8 |
|  | Scholarship | 6 | 1.5 |
|  | Total | 397 | 100.0 |
| Do you do any part-time job, apart from studying? | No | 294 | 73.5 |
|  | Yes | 106 | 26.5 |
|  | Total | 400 | 100.0 |
| If yes, about how many hours do you spend on this job weekly? | Less than 15 hours | 66 | 63.5 |
|  | 15 to 20 hours | 23 | 22.1 |
|  | 21 to 25 hours | 3 | 2.9 |
|  | 26 to 30 hours | 4 | 3.8 |
|  | more than 30 hours | 8 | 7.7 |
|  | Total | 104 | 100.0 |
| Average monthly income (in naira) | Less than 20,000 | 66 | 22.7 |
|  | 20,000 - 49,000 | 155 | 53.3 |
|  | 50,000 - 99,000 | 56 | 19.2 |
|  | 100,000 or more | 14 | 4.8 |
|  | Total | 291 | 100.0 |

Figure 2 shows the prevalence of eating disorders amongst participants – **which is 2.9%.**

**Figure 2:**
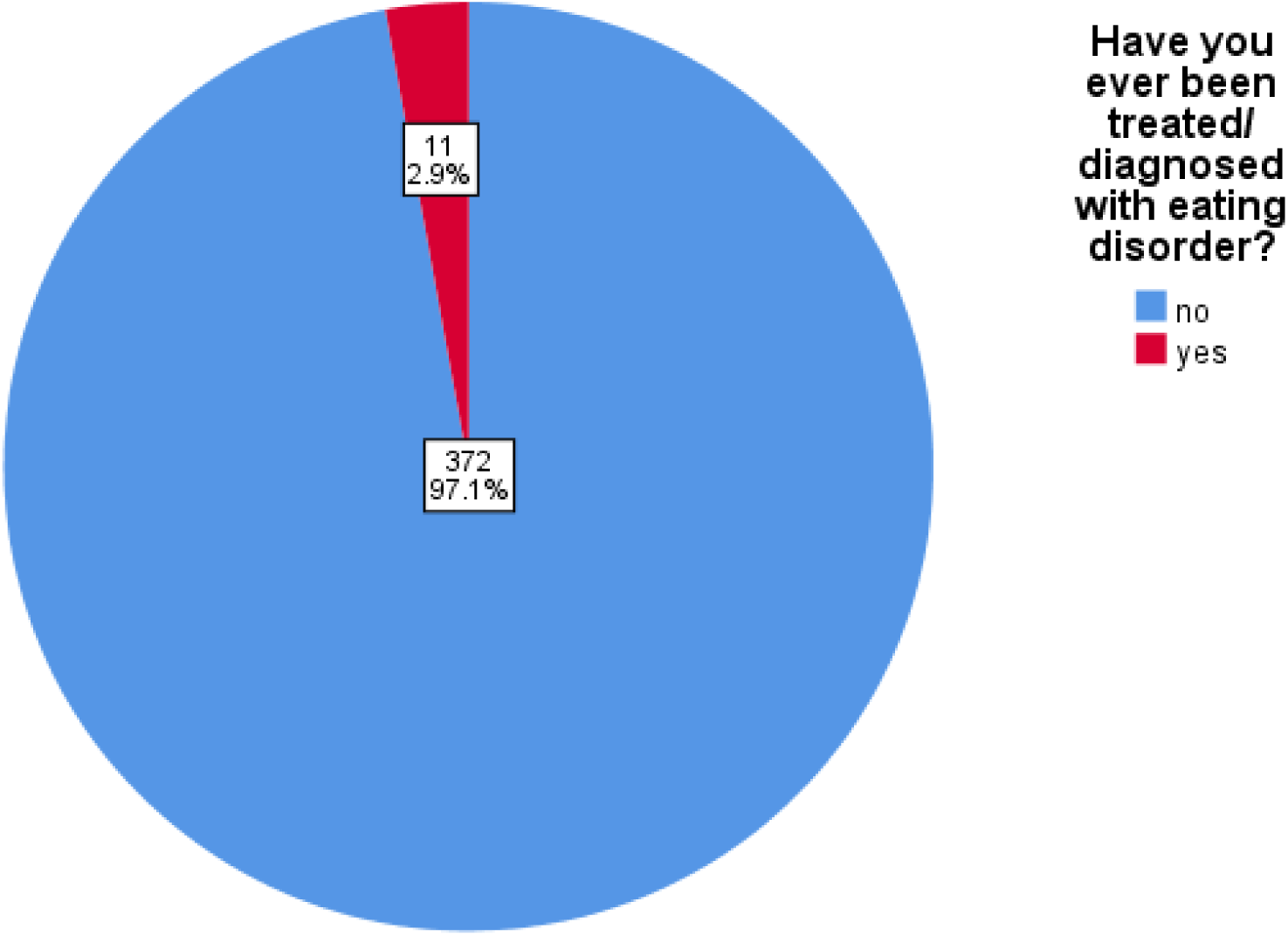
A pie-chart showing prevalence of eating disorders amongst participants (N = 383)

### RISK FACTORS OF EATING DISORDERS AMONGST PARTICIPANTS

**Table 5.**
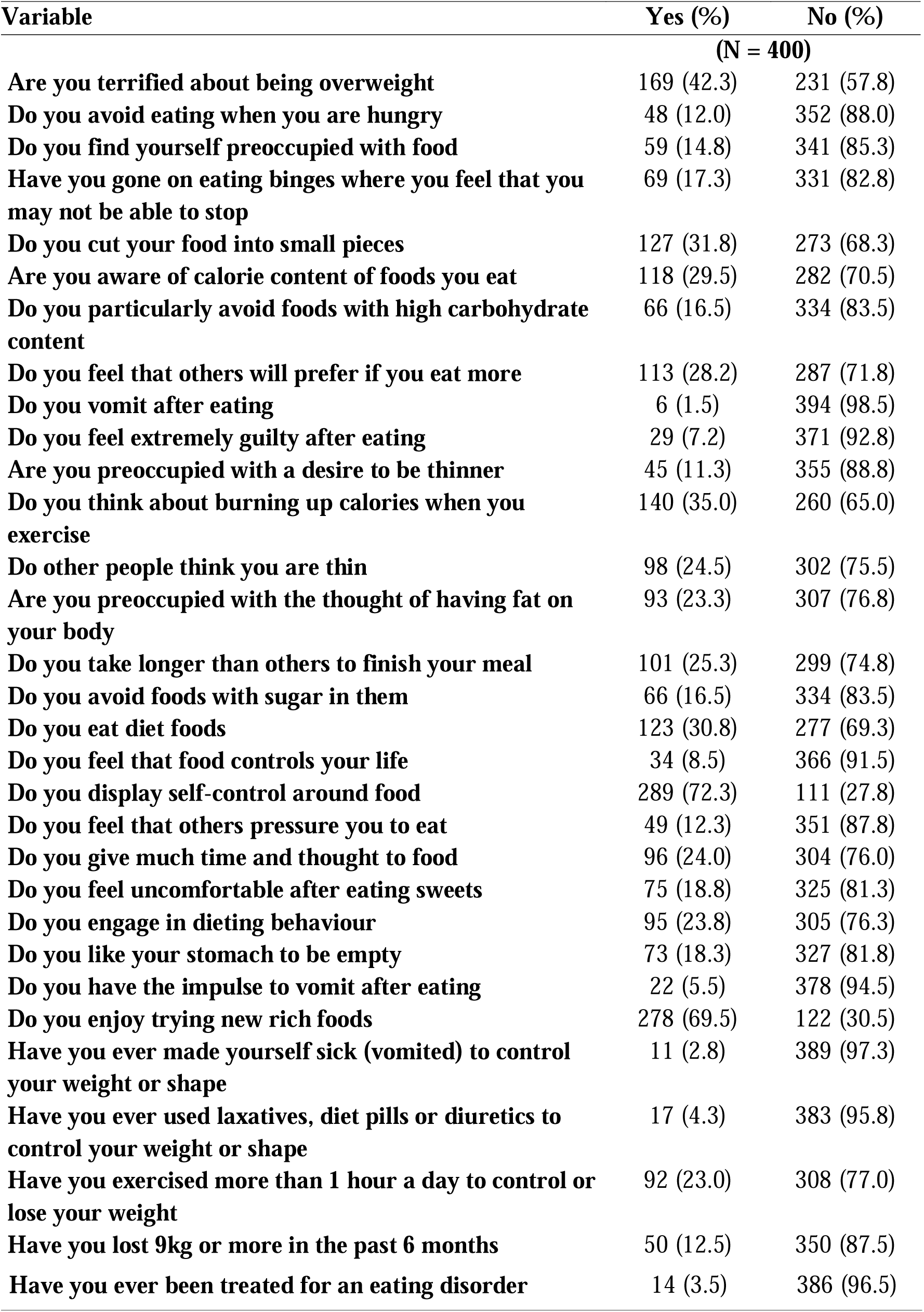
shows risk factors of eating disorders amongst participants.

**Table 4.**
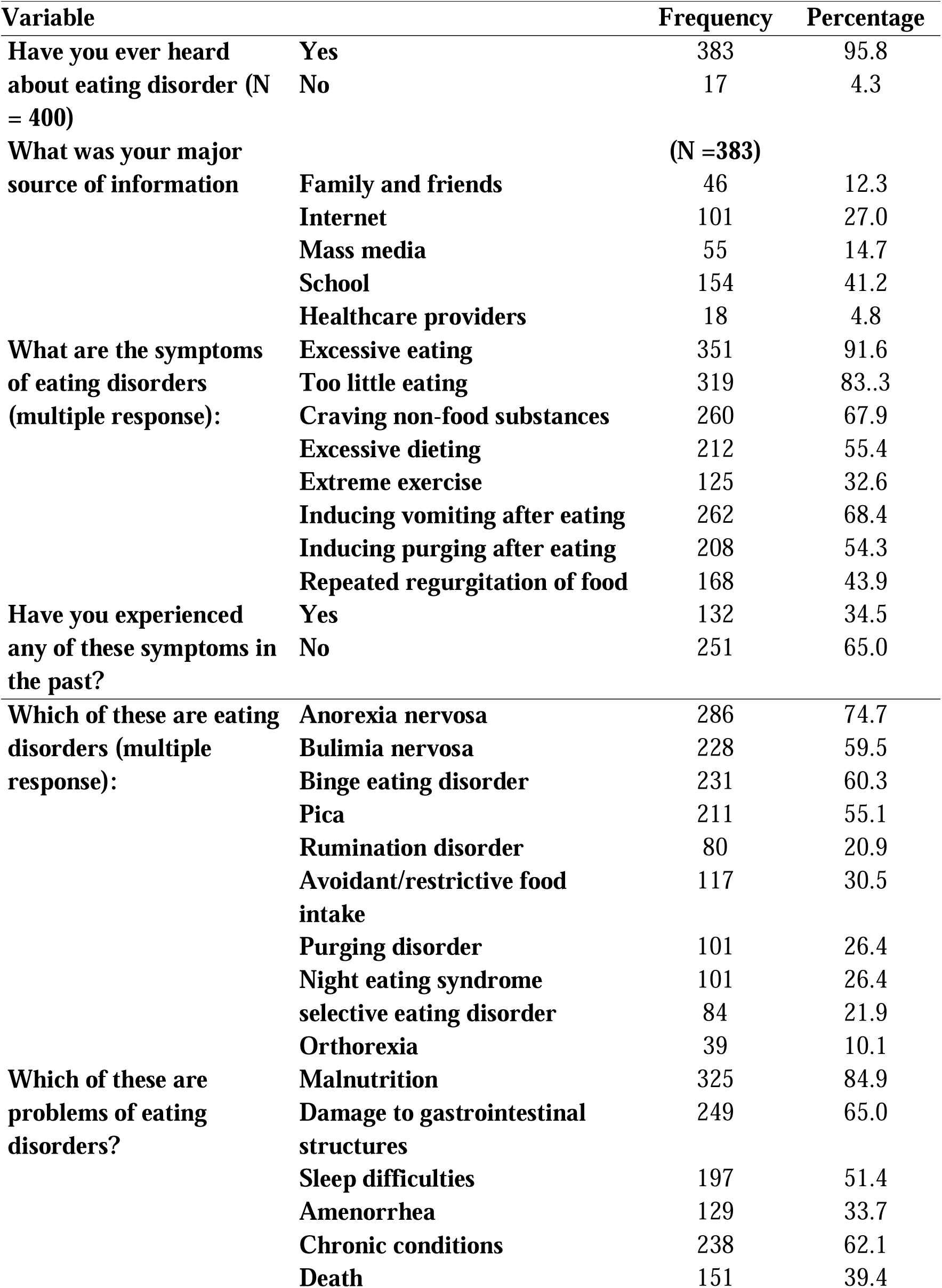
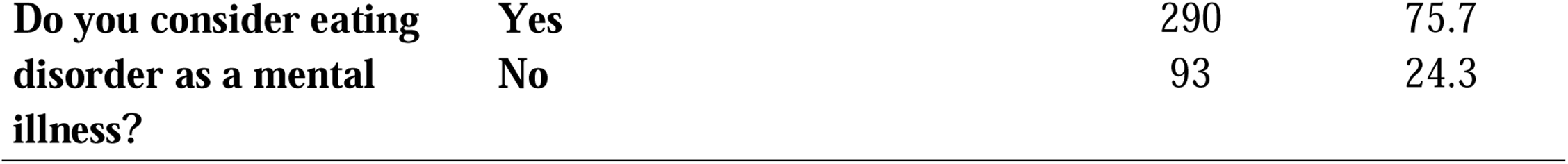
shows participants’ knowledge of eating disorder.

**Figure 3:**
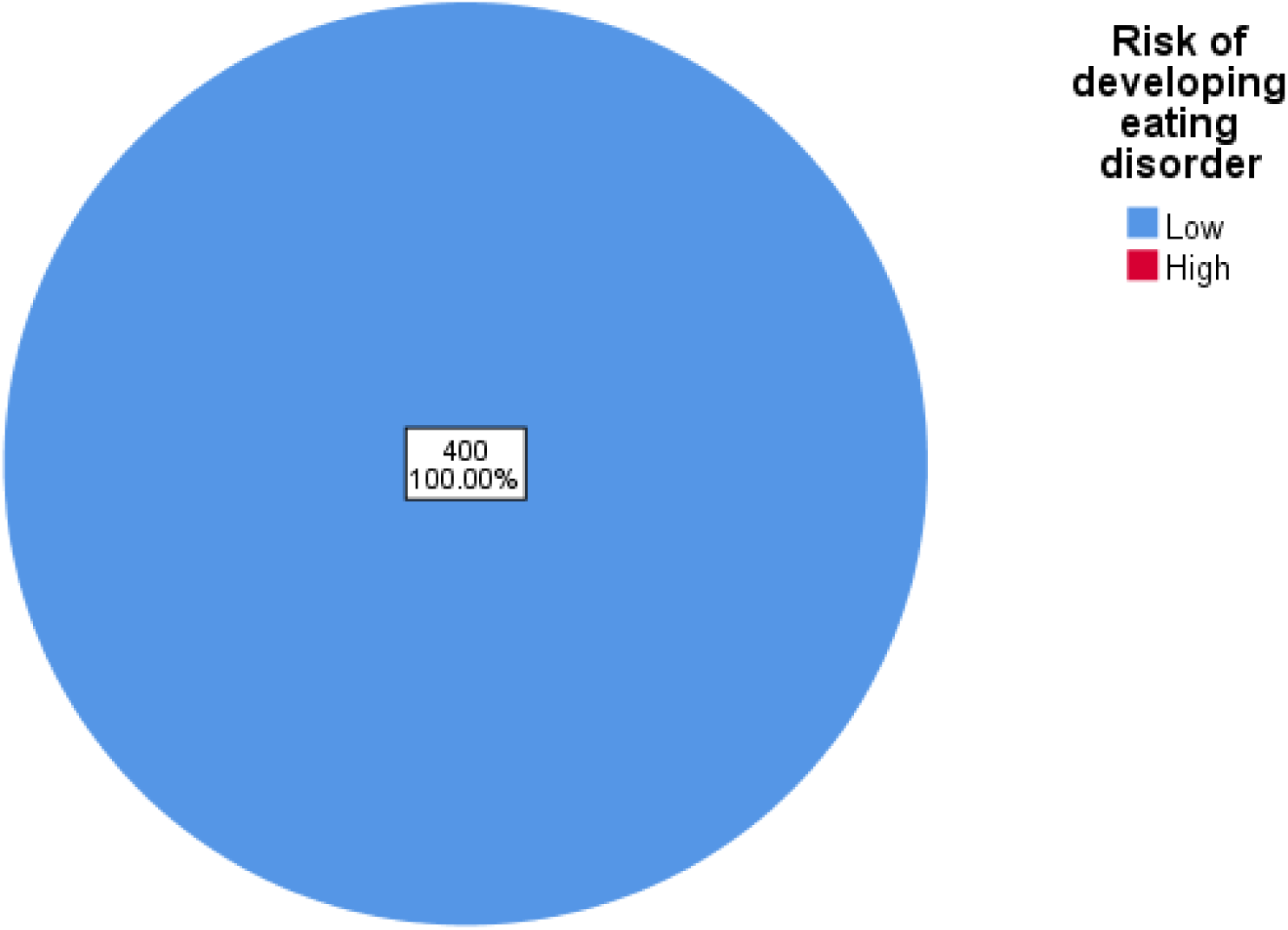
A pie chart showing risk assessment score for developing eating disorder amongst participants. All participants had low risk.

### RELATIONSHIP BETWEEN DIAGNOSIS OF EATING DISORDER AND SOME VARIABLES

|  |  | Have you ever been treated/diagnosed of eating disorder? |  | t / X<sup>2</sup> | p-Value |
| --- | --- | --- | --- | --- | --- |
| Variable |  | No | Yes |  |  |
| Age | Mean | 23.6 | 23.0 | **0.830** | **0.407** |
| BMI | Mean | 23.6 | 30.7 | **4.124** | **0.000\*** |
| Gender | Male | 193 | 7 | **0.139** | **0.709** |
|  | Female | 172 | 5 |  |  |
| Level of Study | 100 - 200 | 9 | 0 | **3.961** | **0.138** |
|  | 300 - 400 | 122 | 7 |  |  |
|  | 500 - 600 | 257 | 5 |  |  |
| Do you smoke | No | 358 | 11 | **0.006** | **0.939** |
|  | Yes | 30 | 1 |  |  |
| Do you take alcohol | No | 215 | 8 | **0.598** | **0.439** |
|  | Yes | 173 | 4 |  |  |
| Average monthly income (in naira) | less than 20,000 | 61 | 5 | **10.062** | **0.018\*** |
|  | 20,000 - 49,000 | 153 | 2 |  |  |
|  | 50,000 - 99,000 | 56 | 0 |  |  |
|  | 100,000 or more | 14 | 0 |  |  |
| How many hours do you sleep every day | 4 – 5 | 92 | 5 | **3.465** | **0.177** |
|  | 6 – 7 | 233 | 6 |  |  |
|  | > 7 | 51 | 0 |  |  |
| Do you skip meals | No | 118 | 0 | **5.177** | **0.023\*** |
|  | Yes | 270 | 12 |  |  |
| How would you grade your academic stress | Mild | 37 | 1 | **0.826** | **0.843** |
|  | moderate | 181 | 7 |  |  |
|  | Severe | 101 | 3 |  |  |
|  | life-threatening | 64 | 1 |  |  |

t = Independent samples Test; X<sup>2</sup> = Chi-square; \* test statistics is significant at 0.05 level.

There is a statistically significant relationship between diagnosis of eating disorder and BMI, average income of participants, as well as history of skipping meals amongst participants.

### PREVALENCE OF PEPTIC ULCER DISEASE AMONG PARTICIPANTS

**Figure 4:**
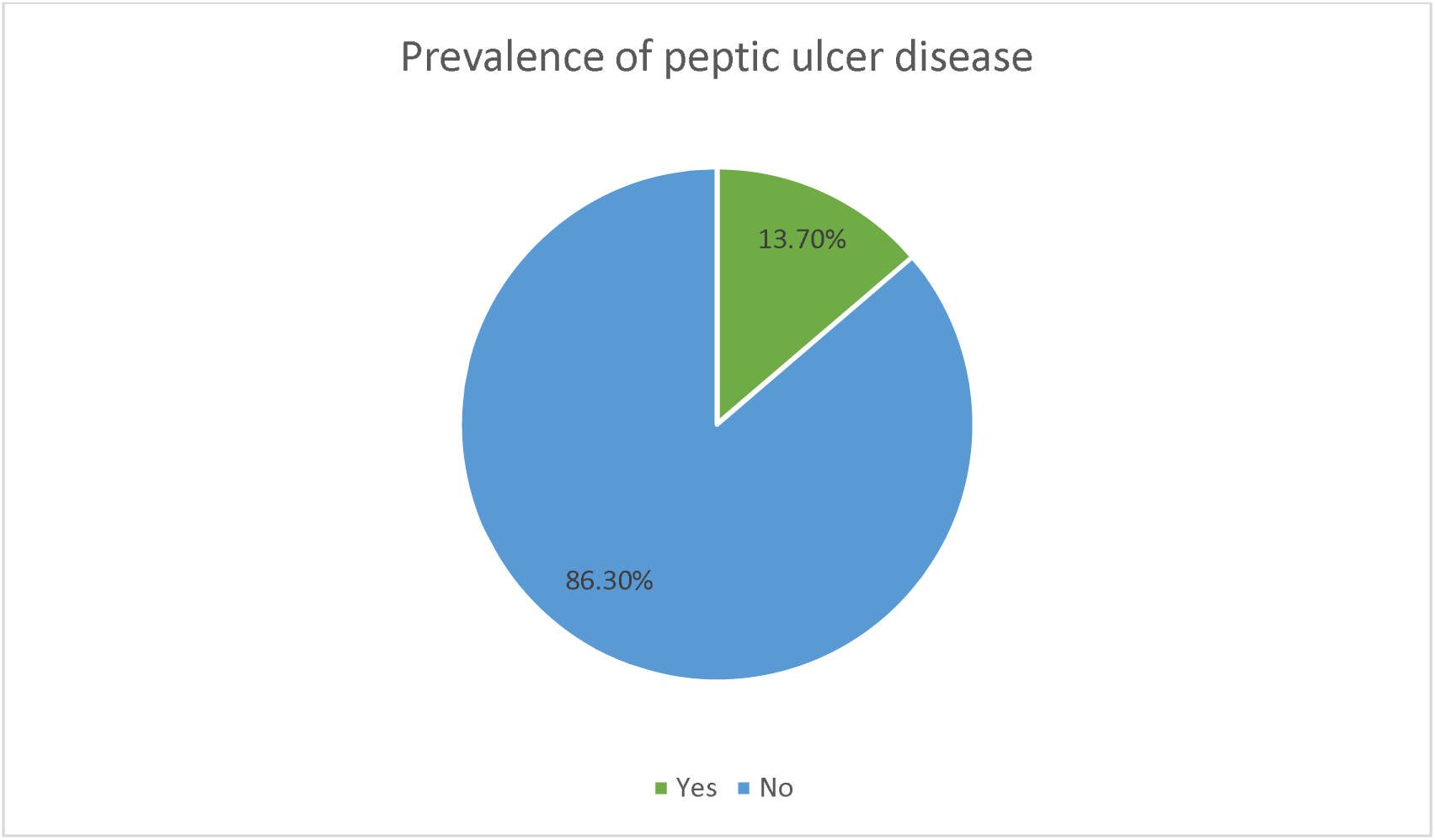
A pie chart showing the prevalence of peptic ulcer disease among participants (N = 380)

### 4.8 RISK FACTORS OF PEPTIC ULCER DISEASE AMONGST PARTICIPANTS

**Table 7.** shows risk factors of peptic ulcer disease amongst participants.

| Variable |  | Frequency<br>(N = 400) | Percentage |
| --- | --- | --- | --- |
| Have you been managed with NSAIDs before | Yes | 224 | 56.0 |
|  | No | 176 | 44.0 |
| If yes, how long |  | (N = 224) |  |
|  | Less than 1 month | 187 | 83.5 |
|  | 1 - 2 months | 15 | 6.7 |
|  | 3 - 6 months | 2 | 0.4 |
|  | Greater than 6 months | 21 | 9.4 |
| How frequently do you take NSAIDs | Always | 2 | 0.9 |
|  | Often | 18 | 8.0 |
|  | Sometimes | 64 | 28.6 |
|  | Rarely | 136 | 60.7 |
|  | Never | 4 | 1.8 |
| Do you take any of the following (Multiple Response) |  | (N = 400) |  |
|  | Coffee | 174 | 43.5 |
|  | Chocolate | 267 | 66.8 |
|  | Caffeinated drinks | 267 | 66.8 |
|  | Spicy foods | 259 | 64.8 |
|  | Alcohol | 159 | 39.8 |
|  | Green tea | 46 | 11.5 |
|  | Bitter kola | 36 | 9.0 |
|  | Cigarettes | 4 | 1.0 |
| How often do you take them | Always | 18 | 4.5 |
|  | Often | 83 | 20.8 |
|  | Sometimes | 202 | 50.5 |
|  | Rarely | 87 | 21.8 |
|  | Never | 10 | 2.5 |

### 4.9 OTHER RISK FACTORS OF PEPTIC ULCER DISEASE AMONGST PARTICIPANTS

**Table 8.** shows other risk factors of peptic ulcer disease amongst participants.

| Variable |  | Frequency<br>(N = 400) | Percentage |
| --- | --- | --- | --- |
| Do you take herbal concoctions | Yes | 16 | 4.0 |
|  | No | 384 | 96.0 |
| What is your blood group | O | 231 | 57.8 |
|  | A | 93 | 23.3 |
|  | B | 46 | 11.5 |
|  | AB | 12 | 3.0 |
|  | Don't know | 18 | 4.5 |
| Do you have a family history of PUD | Yes | 110 | 27.5 |
|  | No | 290 | 72.5 |
| Do you skip meals | Yes | 282 | 70.5 |
|  | No | 118 | 29.5 |
| If yes, how frequently (N = 282) | Always | 32 | 11.3 |
|  | Often | 88 | 31.2 |
|  | Sometimes | 138 | 48.9 |
|  | Rarely | 22 | 7.8 |
|  | Never | 2 | 0.7 |
| How many hours do you sleep every day? | 4 – 5 | 97 | 25.1 |
|  | 6 – 7 | 239 | 61.8 |
|  | > 7 | 51 | 13.2 |
|  | Total | 387 | 100.0 |
| How many hours do you study every day? | 2 - 4 hours | 310 | 80.1 |
|  | 5 - 7 hours | 50 | 12.9 |
|  | 8 - 10 hours | 22 | 5.7 |
|  | More than 10 hours | 5 | 1.3 |
|  | Total | 387 | 100.0 |
| How would you grade your academic stress? | Mild | 38 | 9.6 |
|  | Moderate | 188 | 47.6 |
|  | Severe | 104 | 26.3 |
|  | life-threatening | 65 | 16.5 |
|  | Total | 395 | 100.0 |
| Did this academic stress increase or decrease as you go higher in class? | Increase in stress | 319 | 81.0 |
|  | Decrease in stress | 36 | 9.1 |
|  | Remained the same (no change) | 39 | 9.9 |
|  | Total | 394 | 100.0 |

**Figure 5:**
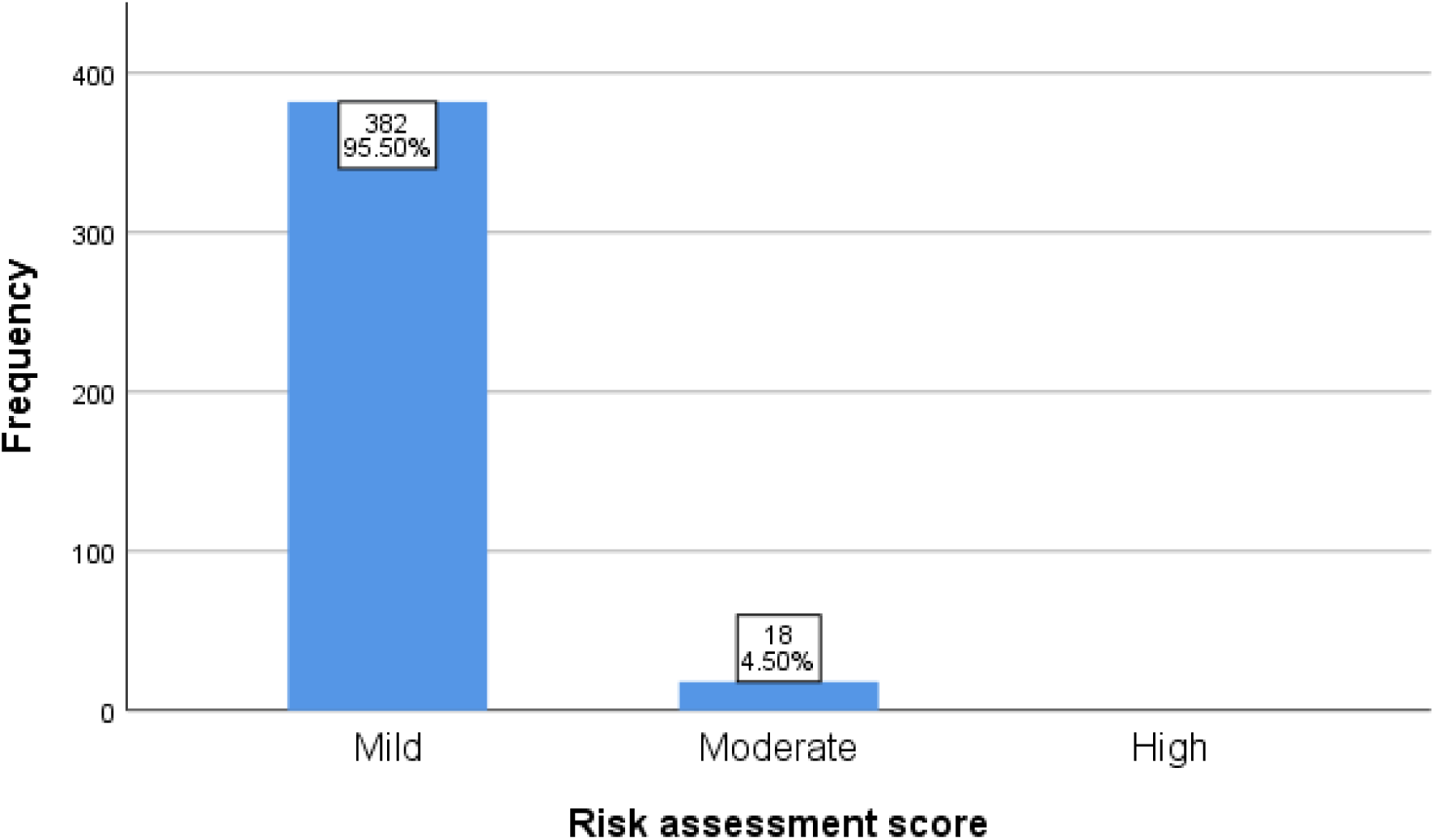
showing the risk assessment score of participants for Peptic Ulcer Disease. Majority (95.5%) of the participants had mild risk assessment score.

### 4.10 PARTICIPANTS KNOWLEDGE OF PEPTIC ULCER DISEASE

**Table 6.**
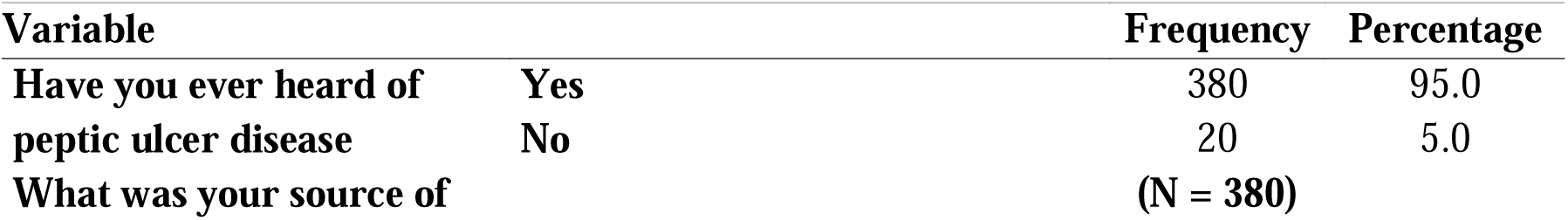
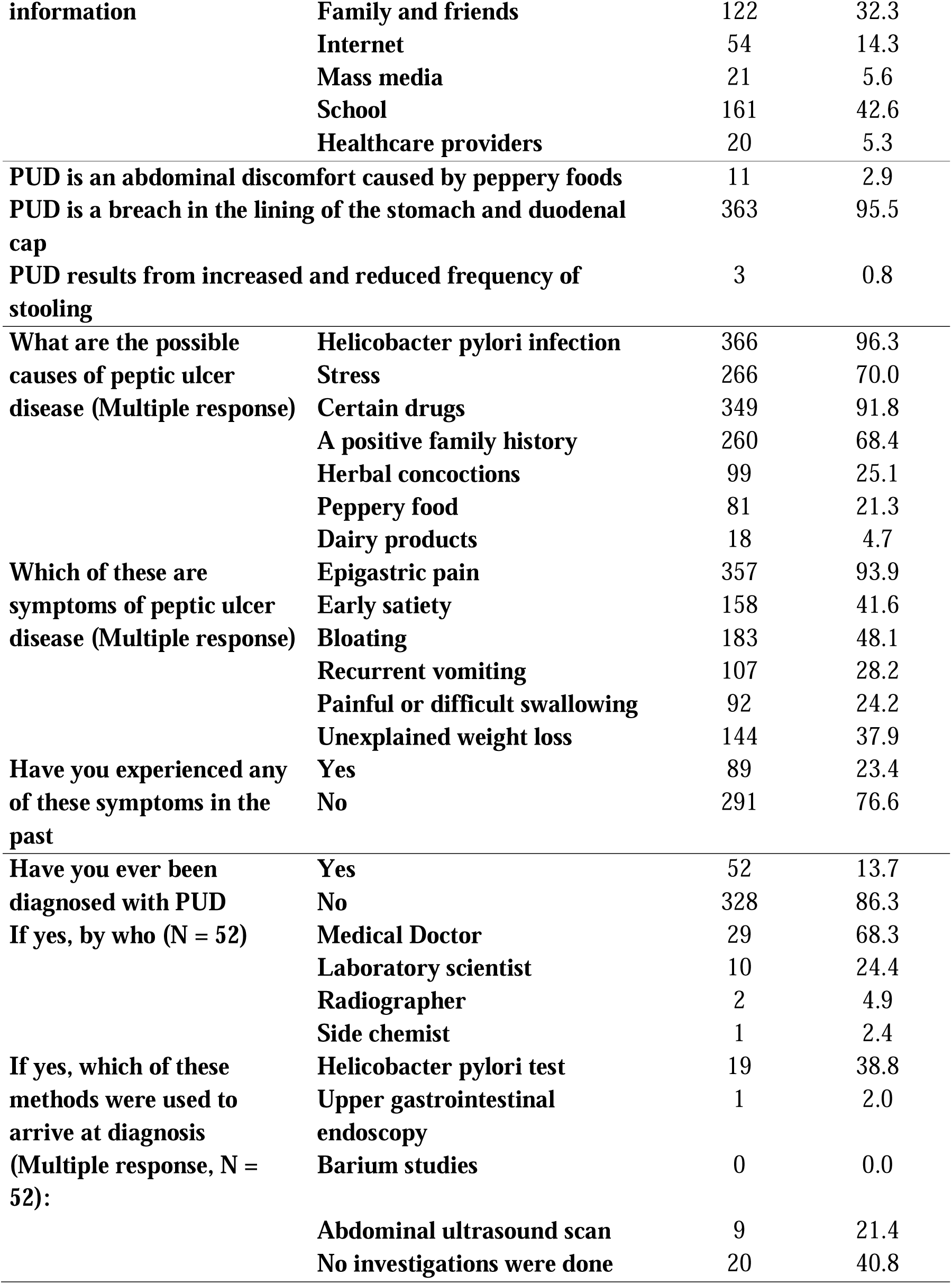
shows participants’ knowledge of peptic ulcer disease.

**Figure 6:**
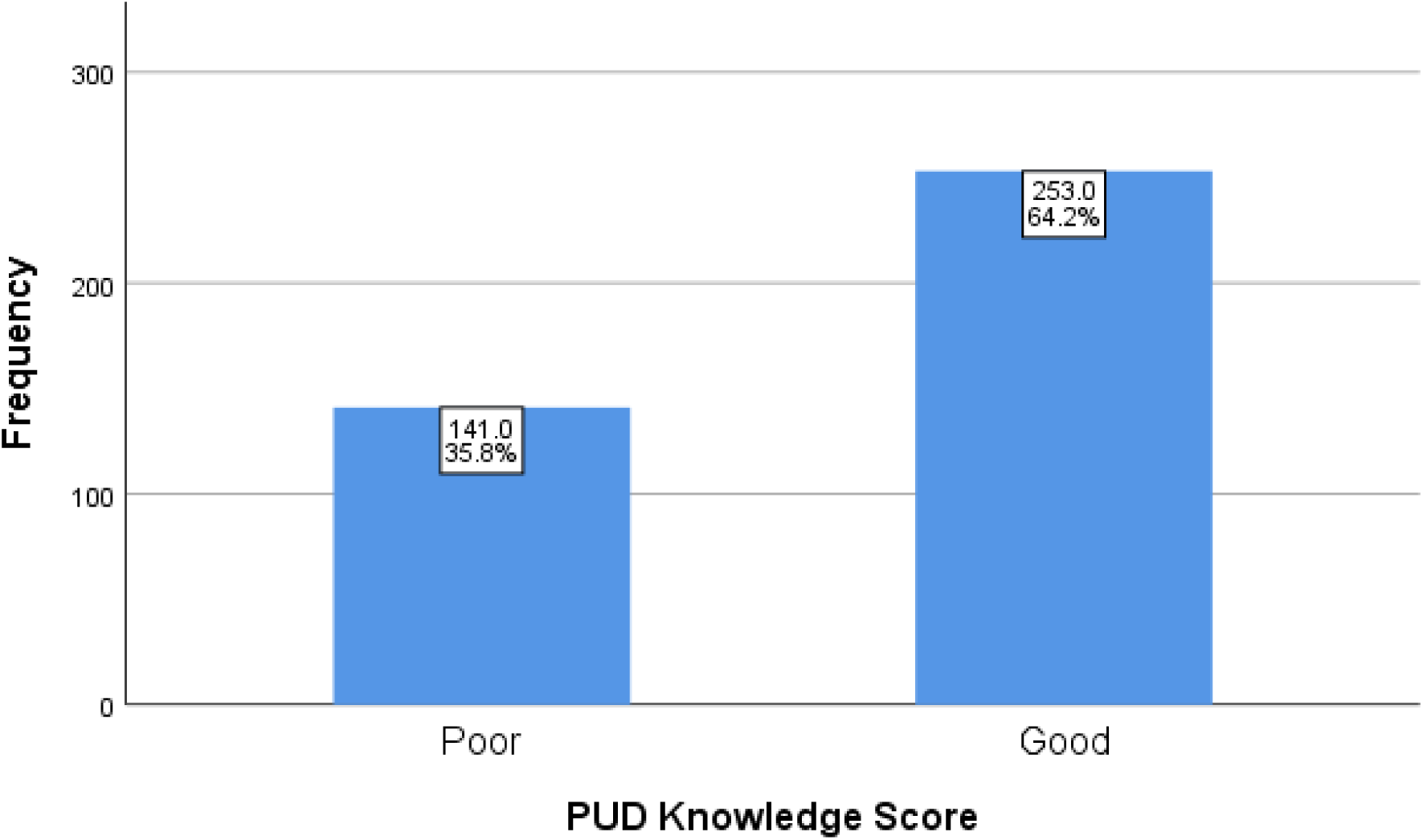
A simple bar chart showing knowledge score of participants about PUD (N = 394)

### RELATIONSHIP BETWEEN PRESENCE OF PUD AND SOME VARIABLES

| Variable |  | Have you ever been<br>diagnosed with PUD? |  | t / X<sup>2</sup> | p-Value |
| --- | --- | --- | --- | --- | --- |
|  |  | No | Yes |  |  |
| Age | Mean | 23.51 | 23.78 | **0.797** | **0.426** |
| BMI | Mean | 23.3 | 23.6 | **0.215** | **0.830** |
| Gender | Male | 176 | 24 | **0.590** | **0.442** |
|  | Female | 151 | 26 |  |  |
| Level of Study | 100 - 200 | 8 | 1 | **0.068** | **0.996** |
|  | 300 - 400 | 112 | 17 |  |  |
|  | 500 - 600 | 226 | 36 |  |  |
| Do you smoke | No | 327 | 42 | **18.289** | **0.000\*** |
|  | Yes | 19 | 12 |  |  |
| Do you take alcohol | No | 200 | 23 | **4.381** | **0.036\*** |
|  | Yes | 146 | 31 |  |  |
| Average monthly<br>income (in naira) | less than<br>20,000 | 54 | 12 | **2.541** | **0.468** |
|  | 20,000 -<br>49,000 | 137 | 18 |  |  |
|  | 50,000 -<br>99,000 | 50 | 6 |  |  |
|  | 100,000 or<br>more | 13 | 1 |  |  |
| Have you been<br>managed with<br>NSAIDs before | No | 157 | 19 | **1.969** | **0.161** |
|  | Yes | 189 | 35 |  |  |
| What is your blood<br>group | O | 196 | 35 | **4.898** | **0.298** |
|  | A | 85 | 8 |  |  |
|  | B | 39 | 7 |  |  |
|  | AB | 9 | 3 |  |  |
|  | Unknown | 17 | 1 |  |  |
| Do you take herbal<br>concoctions | No | 335 | 49 | **4.497** | **0.034\*** |
|  | Yes | 11 | 5 |  |  |
| Do you have a<br>family history of<br>PUD | No | 256 | 34 | **2.848** | **0.091** |
|  | Yes | 90 | 20 |  |  |
| Do you skip meals | No | 105 | 13 | **0.884** | **0.347** |
|  | Yes | 241 | 41 |  |  |
| How would you<br>grade your academic<br>stress | Mild | 32 | 6 | **4.063** | **0.255** |
|  | Moderate | 169 | 19 |  |  |
|  | Severe | 87 | 17 |  |  |
|  | **Life-threatening** | 53 | 12 |  |  |
| **How many persons are you in a room** | **1 – 2** | 224 | 33 | **0.288** | **0.866** |
|  | **3 – 4** | 79 | 14 |  |  |
|  | **More than 4** | 32 | 5 |  |  |
| **what is your method of sewage disposal** | **Water closet** | 312 | 46 | **4.013** | **0.134** |
|  | **Open defecation** | 1 | 1 |  |  |
|  | **Bucket toilet** | 26 | 7 |  |  |
| **What is your source of drinking water?** | **Borehole** | 15 | 0 | **5.634** | **0.068** |
|  | **Sachet water** | 324 | 51 |  |  |
|  | **Bottled water** | 6 | 3 |  |  |

t = Independent samples Test; $X^2$ = Chi-square; \* test statistics is significant at 0.05 level.

## DISCUSSION

The mean age of our participants was 24 years (SD = 2.4) which is closely related to a Bangladesh study with a mean age of 21 years. ^15^ Most (53.1%) were males, Christians (98.7%) and of Igbo ethnicity (98.4%) which could be due to the location of study. Majority (93.6%) reside in the hostels provided by the school and 66.4% live alone or have a roommate, 24% are 3 or 4 in a room while 9.6% have more than four occupants sharing a room. From our study, the prevalence of overcrowding among clinical students is 9.6% which disagrees with other studies ^16–18^ where overcrowding was considered a major stressor among students in Nigeria. This could be because when compared to others, the university provided better welfare for clinical students.

The calculated body mass index shows that majority (69.9%) had normal weight, 18.5% were overweight, 7.3% were obese while 4.6% were underweight. This is similar to a study by Agu et al ^19^ in which the mean BMI was 23.2 +/- 2.5 and another study by Emmanuel et al among nursing students in which 73.3% had a normal weight. ^20^ Most clinical students are aware of the complications of unhealthy weight as this makes up a significant part of their curriculum and they also interface with patients with such complications on a regular basis.

### Eating disorders

Our study shows a 2.9% prevalence rate of eating disorders among our participants which is significantly lower than reported rates of 14.1% by Oyewumi and Kazarian, ^21^ and 10.4% by Jahrami et al. ^22^ Eating disorders our participants have been diagnosed with are anorexia nervosa (0.3%), purging disorder (0.8%) and rumination disorder (0.3%). Two participants (0.6%) wrongly reported gastroenteritis and gastroesophageal reflux disease (GERD) as eating disorders. In Ile-Ife, the high prevalence rates of 20.5% and 17.0% for anorexia nervosa and bulimia nervosa ^23^ are similar to studies in Japan where the prevalence rates of anorexia nervosa and bulimia nervosa were 17.10% and 5.79% respectively. ^15^ In our study, males had a higher prevalence of eating disorders (1.7%) than females (1.2%) which does not agree with other studies in which females have a higher risk of developing eating disorders than males. ^25–28^ Although our study revealed that all participants had a low risk of eating disorders, Jahrami et al reported that medical students were at higher risk of developing eating disorders than the general population. ^22^ The low prevalence of eating disorders from our study could be because our respondents are clinical students who are knowledge about its adverse effects and have therefore, adopted healthy eating habits.

89.7% reported that their education was sponsored by their parents and 53.3% received an average monthly income between 20,000 naira and 49,000 naira. However, about 26.5% worked part-time jobs to cater for their education and 22.1% received less than 20,000 naira from their sponsors. There was a statistically significant relationship between average income of participants **(p=0.018)** and skipping meals **(p=0.023)** in the diagnosis of eating disorders. Some studies linked financial difficulties among students to eating disorders ^26,29^ while Onyeke et al noted that no relationship existed between eating disorders and financial problems. ^30^ Our result agrees with others that skipping meals was a strong factor in developing eating disorders. ^26,31^ The high BMI (mean BMI of 30.7) among our respondents who have been diagnosed with eating disorders suggest that eating disorders was strongly associated with high BMI as demonstrated by other studies ^26,32–34^ although, it was not statistically significant. Academic year of study, smoking, sleeping patterns and alcohol consumption have no influence on eating disorders but similar studies suggest that all of these play a role. ^15,35^

95.8% of participants had heard of eating disorders with school being their major source of information (41.2%) which is similar to a study in Babcock ^36^ where 94.8% were aware of eating disorders. Most respondents correctly identified too little eating (83.3%), inducing vomiting after eating (68.4%), craving non-food substances (67.9%), excessive dieting (55.4%), inducing purging after eating (54.3%), repeated regurgitation of food (43.9%), and extreme exercise (32.6%) as symptoms of eating disorders. 75.7% correctly identified eating disorder as a mental illness while there was a poor knowledge of the different types of eating disorders. Anorexia nervosa (74.7%), binge eating disorder (60.3%), bulimia nervosa (59.5%) and pica (55.1%) were the most identified as eating disorders by participants while avoidant/restrictive food intake (30.5%), purging disorder (26.4%), night eating syndrome (26.4%), selective eating disorder (21.9%), rumination disorder (20.9%) and orthorexia (10.1%) were the least known. Although they were knowledgeable about the more common types of eating disorder, they demonstrated a poor knowledge of the less common eating disorder. A study among nursing students in Nepal also reported that 60% of their respondents had inadequate knowledge of eating disorders. ^37^ Most respondents correctly identified the clinical problems of malnutrition (84.9%), damage to gastrointestinal structures (65%), chronic conditions e.g., obesity, heart conditions, type 1 and 2 diabetes mellitus (62.1%), sleep difficulties (51.4%), amenorrhea (33.7%) and in severe cases, death (39.4%) associated with eating disorders and thus, is in contrast to only 7.5% in a study in Italy. ^38^

### Peptic ulcer disease

The prevalence of peptic ulcer disease among our participants was 13.7% which is close to a global prevalence rate of 5 to 10%. ^39^ The diagnosis of peptic ulcer disease among our respondents was made by a medical doctor in 86.3% of cases with most of the diagnosis investigated by Helicobacter pylori test (38.8%) and abdominal ultrasound scan (21.4%). Despite being the most reliable investigation of choice, endoscopy was the least employed method (2%) for diagnosing peptic ulcer disease among our respondents, which might be because it is an invasive procedure. There is a relatively low prevalence of Helicobacter pylori among our respondents when compared to 87.7% to 89.7% in the general Nigerian population ^40–42^ and 41.3% among students in Ethiopia. ^34^ A study in Nigeria attributes a 27% positivity between Helicobacter pylori infection and peptic ulcer disease which is somewhat similar to the findings from our study. ^43^ However, there is a risk of recall bias as these were subjectively reported by participants in the study.

Using the risk assessment score, 95.5% and 18% of participants had a mild and moderate risk factor of PUD respectively. This could be because majority (94%) sourced their drinking water from filtered water packaged in sachets and sewage disposal was mostly through water closet (91.1%). Good sources of drinking water and proper sewage disposal reduces their risk of acquiring the H. pylori bacteria which is notorious for its orofecal mode of transmission among participants. There is statistically significant relationship between a diagnosis of PUD and positive history of smoking **(p=0.000)**, intake of alcohol **(p=0.036)**, and use of herbal concoctions **(p=0.034)**. This corresponds with studies that reported the risk of PUD increased with year of study, frequent use of NSAIDs, smoking, prolonged fasting, alcohol intake, H pylori infection and anxiety. ^44–46^ However, unlike these studies, ours showed no relationship between increasing age, year of study or having a blood group O and peptic ulcer disease. Most respondents (83.5%) used NSAIDs for less than one month, only 27.5% had a positive family history of PUD, 61.8% slept for 6-7 hours every day, but 72.7% reported that they skipped meals and majority (47.6%) graded their stress as moderate. These could account for the decreased risk factors and hence, low prevalence rate of peptic ulcer disease among our participants.

95.5% correctly defined peptic ulcer disease as a breach in the lining of the stomach and duodenal cap but only 64.2% knew the causes and symptoms of peptic ulcer disease. This agrees with a study in Ghana in which 64% and 53.5% of medical and nursing students respectively had a good knowledge of peptic ulcer disease. ^42^ In India, out of 200 adolescents in pre-university colleges, majority (47.5%) had a moderate level of knowledge regarding peptic ulcer disease while 18.5% and 0.5% had good and excellent knowledge respectively. ^[50]^ However, a study in North-Central Nigeria revealed that very few undergraduate students had a good knowledge of peptic ulcer disease. ^[51]^ At this level in their medical training, we expected the participants to demonstrate a much higher knowledge of peptic ulcer disease than was observed.

## RECOMMENDATIONS

Our study shows that majority of the respondents have heard about eating disorders and peptic ulcer disease but only an average number have accurate information about the types of eating disorders and the clinical problems associated with them. Given their common presentation, more lectures should be introduced into the curriculum of clinical students to ensure the easy identification and treatment of these disease entities. Scholarship opportunities, free meals, student loans and other forms of welfare and financial aid should be made readily available for students to enable them afford/access clean drinking water, good food and reduce the amount of stress they pass through.

Clinical students should be encouraged to participate in sports and other forms of exercise. This will go a long way in keeping their BMI within normal range since high BMI was found to be associated with eating disorders in the study.

Regularly screening clinical students with non-modifiable risk factors for eating disorders and/or peptic ulcer disease such as positive family history, blood group O, etc. to ensure early diagnosis and treatment should be encouraged. To ensure early presentation to clinics, the cost of doctor consultation, tests and other relevant healthcare services should be subsidized or completely waived for clinical students. All of these will further reduce their prevalence and exposure to risk factors of eating disorders and peptic ulcer disease.

## Data Availability

All data produced in the present work are contained in the manuscript

## Notes

### Competing Interest Statement

The authors have declared no competing interest.

### Funding Statement

This study did not receive any funding

### Author Declarations

Health Research and Ethics Committee, University of Nigeria Teaching Hospital (UNTH), Ituku-Ozalla, Enugu State gave ethical approval for this work

